# A Three-Stage Algorithm for Quantification of the MMSE Interlocking Pentagon Areas

**DOI:** 10.1101/2023.09.04.23294134

**Authors:** Namhee Kim, Timothy Truty, S. Duke Han, Moonseong Heo, Aron S. Buchman, David A. Bennett, Shinya Tasaki

## Abstract

The Mini-Mental State Examination (MMSE) is a widely employed assessment tool for measuring the severity of cognitive impairment. Among the MMSE items, the pentagon copying test (PCT) requires participants to accurately replicate a sample of two interlocking pentagons. While the PCT are traditionally scored on a binary scale, there has been limited developments of granular scoring scale to assess task performance. In this paper, we present a novel three-stage algorithm, called Quantification of Interlocking Pentagons (QIP), which quantifies PCT performance by computing the areas of individual pentagons and their intersection areas, and a balance ratio between the areas of the two individual pentagons. The three stages of the QIP algorithm include: (1) detection of line segments, (2) unraveling of the interlocking pentagons, and (3) quantification of areas. The QIP algorithm was applied to 497 cases from 84 participants. Analysis of the quantified data revealed a significant inverse relationship between age and balance ratio between two pentagon areas (beta = -0.49, p = 0.0033), indicating that older age was associated with a smaller balance ratio. The QIP algorithm enhanced the scoring of performance in the PCT. It can serve as a useful tool for granular level scoring of PCT.

## Introduction

The Mini-Mental State Examination (MMSE), introduced by Folstein et al^1^. in 1975, is a widely utilized 30-point evaluation for assessing delirium and cognitive impairment in clinical and research settings. One of the items in the MMSE is the pentagon copying test (PCT), which requires participants to replicate a sample of two interlocking pentagons on a paper. This test is used to measure visual-spatial abilities and provides psychomotor information about visual-spatial construction, fine motor coordination, and attention to detail.

The traditional scoring of the PCT in the MMSE is binary; (correct=1) if their drawing has ten angles and two pentagons that intersect. A study was conducted to explore scoring at a more detailed level, using qualitative measures, where cases with a score of zero were further categorized into five classes based on the degree of deviation from the interlocking pentagon sample used for testing ^2^. A composite score, which was the sum of five domain scores based on the number of angles, distance/intersection between two pentagons, closure/opening of the image contour, rotation, and closing-in was proposed^3^. To enhance efficiency and objectivity, automatic scoring approaches utilizing Deep Learning (DL) techniques, such as U-Net and convolutional neural networks, have been proposed ^4-6^. A study developed DL method for a mobile application, which employed a convolutional network known as U-net along with mobile sensor data^4^. Another study adopted a convolutional neural network utilizing an object detection model to generate an automatic standard binary score^5^. While two preceding studies focused on training deep learning models to automate two well-established scoring systems, a study devised a deep learning approach to explore an optimal scoring method for the interlocking pentagons test’s correlation with cognition^6^. Additionally, eight crucial drawing characteristics were identified through simulations using synthetic interlocking pentagon images including differences in sizes between two pentagons, overall pentagon size, and distance between the pentagons.

In this study, we selected three attributes as highlighted in Tasaki et al.^6^, and conducted quantification for the targeted attributes. We hypothesized that individuals with impaired visual construction skills, particularly older adults, may encounter difficulties accurately reproducing the sample interlocking pentagons. This difficulty could be reflected in several ways, including a smaller total area, a reduced proportion of intersecting area, and an imbalance in the area between the two pentagons.

We developed a three-stage algorithm called Quantification of Interlocking Pentagon (QIP) to quantify specific metrics, including the areas of individual pentagons, the proportion of intersecting areas, and the balance ratios between the pentagon areas. To this end, the QIP algorithm specifically targeted PCTs that met the criteria for a “correct” condition (score=1) based on the standard binary scoring method. The algorithm comprises line segment detection, unraveling of the interlocking pentagons, and quantification of relevant areas. Figure 1 provides a visual representation of the QIP algorithm process. We applied the algorithm to a subset of 90 randomly selected participants from the Rush Memory and Aging Project (MAP), an ongoing cohort study investigating aging and dementia while cognitive status and MMSE scores, including the binary PCT scores, were blinded. We present the results of our study in Section 2, followed by a discussion in Section 3. Additionally, we provide detailed information about the materials, methods and statistical analysis used in Section 4. Supplementary Materials are also included to offer descriptions of the QIP algorithm components.

## Results

### Participants information

We selected 557 PCTs from 90 participants randomly from MAP, an ongoing cohort study investigating aging and dementia while cognitive status and MMSE scores, including the binary PCT scores, were blinded. Average age at baseline was 82 years old (SD 6.2), education 15 years (SD 2.8), men 25 (27.8%), non-Latino White 89 (98.9%). Participants had follow-up visits on average 6.6 years (SD= 3.4). Demographic data of participants are described in Table 1.

**Table 1.** Baseline demographic distribution and total number of visits from all participants (n=90)

| Variable | Mean | SD or % | Min | 25<sup>th</sup> | 75<sup>th</sup> | Max |
| --- | --- | --- | --- | --- | --- | --- |
| Age at baseline | 81.9 | 6.2 | 68.1 | 77.2 | 85.6 | 99.8 |
| Education | 15.0 | 2.8 | 6.0 | 13.0 | 16.0 | 23.0 |
| Male (n, %) | 25 | 27.8 | - | - | - | - |
| Non-Latino White (n, %) | 89 | 98.9 | - | - | - | - |
| N of total visits | 6.1 | 3.5 | 1 | 4 | 9 | 13 |

### PCT cases incorporated for quantification employing the QIP algorithm

Of 557 PCTs, 56 were scored zero based on the standard binary scoring and were excluded from quantification using the QIP algorithm. The excluded images exhibited arbitrary shapes, including single pentagons, two interlocking rectangles, no intersection between the two pentagons, or images with unintentional movements. Supplementary Figure 1 presents examples of these excluded cases. A total of 501 PCTs with a score of 1 from 85 participants were included for quantification using the QIP algorithm. We evaluated the QIP algorithm’s performances with the original images manually. Among the 501 PCTs, the algorithm failed in quantification for 4 PCTs. These challenges were attributed to very low scan quality (n=3), where lines were not distinguishable from the background that led to failure of detection of lines from the image at Stage 1 of the QIP, or highly wavy drawings that led to failure of accurate estimation of pentagon areas at Stage 3 of the QIP (n=1). Consequently, a total of 497 PCTs from 84 participants were successfully quantified.

### Association of three metrics from the QIP with demographic variables

At baseline, we examined the distribution of three metrics obtained from the QIP algorithm. On average, the proportion of intersection to the total area was 9.8% (SD=4.4%). The balance ratio, which represents the ratio of the smaller pentagon to the larger pentagon, was 82.2% (SD=12.2%). The ratio of the total area to the sample interlocking pentagon administered for PCT was 138% (SD=78%) (Table 2). As a reference, the sample interlocking pentagon displayed a proportion of intersection of 6.2% and a balance ratio of 99.0%. We observed a positive correlation between the balance ratio and the proportion of intersecting area (Spearman Correlation=0.274, p=0.0117), indicating that a higher balance ratio was associated with a greater proportion of intersection (Table 3 and Figure 2).

**Table 2.** Summary of PCT quantification at baseline (n=84)

|  | Mean | SD | Min | 25% | 75% | Max |
| --- | --- | --- | --- | --- | --- | --- |
| Total Area (%) | 139 | 78 | 25 | 91 | 162 | 456 |
| Prop. Intersection (%) | 9.8 | 4.4 | 1.7 | 6.5 | 12.7 | 22.5 |
| Balance Ratio (%) | 82.2 | 12.2 | 55.4 | 71.9 | 93.5 | 99.9 |

Note: Total area was divided by the total area of the sample interlocking pentagon administered for PCT.

**Table 3.** Spearman Correlation among three metrics of PCT at baseline (n=84)

|  | Prop. Intersection Area | Balance Ratio |
| --- | --- | --- |
| Total Area | 0.0244 (p=0.8536) | 0.06637 (p= 0.5486) |
| Prop. Intersection Area | 1.0 | 0.27387 (p= 0.0117) |

Figure 3 illustrates the distribution of the three measures (total area, proportion of intersection, and balance ratio) among three age groups at baseline based on tertiles (age <79, 79≤ age <82, age ≥82). Spaghetti plots depicting the longitudinal measures against age are presented in Figure 4. Linear mixed-effects models were employed to analyze the relationship between each longitudinal measure and age at baseline, sex, years of education, lag (in years) since enrollment of the study, and the interaction between lag and the three demographic measures with 497 PCT cases from 84 participants. We assessed the normality assumption of each QIP metric as an outcome of the mixed effects model, and determined that all three metrics were acceptable for normality assumption^7, 8^. The results showed a significant inverse relationship between age and the balance ratio (beta = -0.49, p = 0.0033), indicating that a two-year increase in age at baseline was associated with approximately a 1% decrease in the balance ratio. Furthermore, the balance ratio exhibited a decline over the follow-up visits (beta = -5.44, p = 0.0394), suggesting an annual decrease of approximately 5.4%. Furthermore, we observed significant interaction effects between age and lag (beta = 0.07, p = 0.0372) as well as between sex and lag (beta = -1.06, p = 0.0104), indicating that the associations of balance ratio with age at baseline and sex change over the course of follow-up visits. Additionally, education was found to be associated with the proportion of intersecting area, with every 2-year increase in education being associated with a 1% decrease in the proportion of intersecting area (beta = -0.53, p = 0.0005). However, no significant relationship was found between the total area and any of the three demographic variables. A summary of these results can be found in Table 4.

**Table 4.** Linear mixed effects model with three longitudinal PCT measures.

|  | Effect | Estimate | SE | Pr > \|t\| |
| --- | --- | --- | --- | --- |
| Total Area (%) | Age | 0.62 | 1.20 | 0.6070 |
|  | Sex | 9.92 | 15.84 | 0.5318 |
|  | Education | -1.51 | 2.62 | 0.5643 |
|  | Lag | 5.50 | 15.11 | 0.7170 |
|  | Age*Lag | -0.07 | 0.19 | 0.7115 |
|  | Sex*Lag | 0.06 | 2.34 | 0.9789 |
|  | Education*Lag | -0.23 | 0.37 | 0.5320 |
| Proportion of Intersection (%) | Age | -0.06 | 0.07 | 0.3524 |
|  | Sex | 0.43 | 0.92 | 0.6413 |
|  | Education | -0.53 | 0.15 | 0.0005*** |
|  | Lag | 0.18 | 0.99 | 0.8601 |
|  | Age*Lag | 0.00 | 0.01 | 0.8753 |
|  | Sex*Lag | 0.03 | 0.16 | 0.8706 |
|  | Education*Lag | 0.01 | 0.02 | 0.5745 |
| Balance Ratio (%) | Age | -0.49 | 0.17 | 0.0033** |
|  | Sex | 2.89 | 2.21 | 0.1909 |
|  | education | -0.17 | 0.35 | 0.6265 |
|  | Lag | -5.44 | 2.56 | 0.0370* |
|  | Age*Lag | 0.07 | 0.03 | 0.0372* |
|  | Sex*Lag | -1.06 | 0.41 | 0.0104* |
|  | Education*Lag | 0.07 | 0.06 | 0.2730 |

Age: Age at baseline; \*- p-value <0.05; \*\* p value < 0.01 \*\*\* p value < 0.001

## Discussion

We found that the developed QIP algorithm was successful in quantifying three metrics from the PCTs. The QIP approach provides a more detailed quantification compared to the standard binary scoring, which only assesses the presence of ten angles and the intersection of two pentagons. The QIP algorithm was specifically designed to quantify PCTs that scored correct based on the standard binary scoring, and it provides three metrics: total area, intersecting area, and balance ratio. Among these metrics, we found that balance ratio showed a significant association with age at baseline and duration (in years) since enrollment of the study, proportion of intersection with years of education.

The most common method for quantifying PCT is a straightforward binary scoring system, possibly due to the intricate nature of analyzing interlocking shapes. A more detailed scoring system was proposed, involving the aggregation of five domain scores based on aspects such as angle count, distance/intersection between pentagons, closure/opening of image contours, rotation, and closing-in ^3^. However, this method of quantification could exhibit variation among different raters. To improve efficiency and objectivity, automated scoring approaches employing Deep Learning (DL) techniques have been adopted ^4^. On the other hand, Tasaki et al. ^6^ developed a deep learning model correlating with cognitive scores and introduced eight features, several of which were novel features for PCT characteristics. The QIP algorithm presents a methodology for quantifying three features out of the aforementioned eight proposed by Tasaki et al.^6^. Another noteworthy aspect of the QIP algorithm is its applicability to interlocking polygons beyond pentagons, such as rectangles or triangles as long as objects maintain convexity. However, drawings other than pentagons are indicative of cognitive impairment and receive a score of zero based on standard binary scoring, thus we excluded the application of the QIP algorithm to such cases.

The algorithm encountered difficulties when processing images of poor scan quality, highly waviness or instances of overshooting. In cases of low scan quality, the lines often blended with the background, causing Stage 1 to fail in detecting the lines. Drawings with high waviness or overshooting resulted in the inaccurate calculation of pentagon areas during Stage 3 of the QIP, owing to disparities between the original image and the reconstructed image based on the convex hull approach. It’s worth noting that low scan quality could stem from ongoing neurodegeneration as studied in Tasaki et al.^6^ manifesting as reduced hand strength. On the other hand, overshoots might be reflective of an individual’s distinctive drawing style. For interlocking pentagon images with low scan quality and high waviness, we classified them as failures of the algorithm. Meanwhile, to address overshoots, we introduced an extra manual step between Stage 2 and 3 for images exhibiting this trait. In this step, nodes forming overshoots were removed to rectify the issue.

However, this study has certain limitations. Our developed algorithm is specifically designed to quantify PCTs that meet the requirements for the good condition based on the binary scoring, excluding cases with a score of zero that indicate the severity of cognitive impairment. Subsequent research should emphasize the integration of pre-existing DL methods^4-6^ with the QIP, enabling a comprehensive approach for both classifying and quantifying PCTs. Additionally, the computational complexity of Stage 2 of the QIP algorithm, which incorporates random permutation, warrants further improvements for enhanced efficiency. In this study, we did not examine the associations between the three QIP metrics and cognitive or motor measurements, as our primary focus was on detailing the QIP algorithm. Future studies should investigate how these area metrics are associated with the cognitive and motor abilities of older individuals.

## Materials and Methods

### Ethical statement

All studies were approved by an Institutional Review Board of Rush University Medical Center. Written informed consent was obtained from each participant.

### Materials

The MMSE has been administered for cognitive tests for all cohort studies at Rush Alzheimer’s Disease Center. Common eligibility criteria in both studies included: age > 65 years, absence of known dementia at the time of enrollment, and agreement to annual clinical evaluations. Of over 5000 participants tested with the MMSE at least once, we selected 90 participants blinded with participants’ cognitive status.

### Methods

The PCT was administered on paper, the paper was scanned, and then saved in the portable network graphics (png) format. The digital image was the input for the algorithm. Each digital image was first transformed into a binary image, where pixels are with zero for background and a positive constant for participant’ drawing. We, instead of using a massive number of individual nonzero pixels in each digital image for analysis, utilize line segments each of which is a collection of nonzero pixels.

The algorithm consists of three stages: (1) line segment detection from the image, (2) unraveling of two interlocking pentagons, and (3) quantification of the areas of interest. The first stage of detecting line segment is preceded by edge detection for which we applied the Canny edge detection ^9^ that skeletonizes image. Once the edges are detected, the Hough transformation ^10^ was applied to detect line segments, each of which is characterized by their starting and ending points in Euclidean space. In the second stage, to unravel two interlocking pentagons, we clustered the Hough line segments into individual pentagons based on connectivity matrix using the algorithm we developed. The third stage quantifies areas of individual pentagons using the Monte Carlo integration^11^. Flow of the algorithm is shown in Figure 1. For the purpose of demonstrating the QIP algorithm, we chose three PCTs comprising the sample interlocking pentagon and two additional PCTs from different participants, whose ages ranged from their 70s to their 90s. These three PCTs are illustrated in Supplementary Figure 2.

### Stage 1. Detection of edges and line segments

#### 1.1 The Canny edge detection

The Canny edge detection algorithm ^9^ simplifies the image by keeping only boundaries of the input image. The Canny edge detection algorithm detects pixels in digital image that display a sharp change in intensity, often referred to as edges. Output from the Canny edge detection algorithm is a binary image with a positive value for detected edges. Edges contain shape information and thus are the most important features for image recognition and classification. Keeping only edges in image reduces the amount of data to be processed. Using the image containing only edges from the Canny edge detection algorithm, we identified line segments which resulted in a further reduction of image data into a collection of two end points for each line segment. We demonstrated the Canny edge detection with the three example PCTs in Supplementary Figure 3. Details of the Canny edge detection algorithm are shown in Supplementary Material A.

#### 1.2 The Hough transformation

In two-dimensional Euclidean space, a line can be parameterized by the y-intercept (*α*) and slope (*β*). Although this parameterization is most extensively adopted, it is not able to represent vertical lines for which the slopes are infinity, *i*.*e*., *β* =∞. For this reason, the Hough transformation^10^ is a useful alternative parametrization of lines that enables to detect lines in image using a pair of parameters (*r,θ*) : the angle(*θ*) between a line perpendicular to the line (*l*) and the positive x-axis; the shortest distance (*r*) from the origin (0,0) to the line *l*. Each image from the Canny edge detection algorithm ^9^ was processed by the Hough transformation ^10^ to identify line segments. The pair of parameters (*r,θ*) is referred to as the Hough parameters. That is, a line in the Euclidean space is transformed to a point in the Hough space with (x-axis: *θ*, y-axis: *r*). Any points (*x, y*) on a line in Euclidean space is thus alternatively represented by a pair of (*r,θ*) in the Hough space as follows.

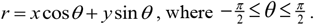

Images were reconstructed with line segments identified from the Hough transformation ^10^. To keep line segments with higher authenticity, three thresholds were applied. Those are threshold for (a) minimal number of points sharing the Hough parameters, (b) minimal length of a line, and (c) minimal gap allowed between two distinct line segments. Two Hough lines with a gap less than the minimal gap were merged. Higher the number of points sharing the same Hough parameters and longer length of a line signify more authenticity of a line. We demonstrated the Hough transformation ^10^ with the three example PCTs in Supplementary Figure 4 and the estimated line segments in Supplementary Figure 5. Details of the Hough transformation ^10^ are shown in Supplementary Material B.

### Stage 2. Separation of two interlocking pentagons

Because two pentagons intersect, quantifying the areas of individual pentagons and their intersection can be complex and challenging. To simplify the computation process, we first aimed to separate the two pentagons. This was done using the line segments acquired in Stage 1, where the line segments were grouped into two individual pentagons. Each line segment detected from the Hough transformation has two endpoints. We call each of the endpoints of a line segment a node so that each line segment has two nodes. It is important to note that assigning the nodes around the centroid, which represents the gravity of all nodes, to their respective pentagons poses a higher level of complexity due to the intersection occurring near the centroid. To address this, we implemented a cut-off distance from the centroid, designating nodes in close proximity as inner nodes, and those further away as outer nodes. We provided further details on cut-off distance in Supplementary Material C. To cluster the nodes into their respective pentagons, we employed different approaches. For the outer nodes, we utilized hierarchical clustering, while for the inner nodes, which required more computational intensity due to their proximity to the centroid, we adopted a hybrid approach combining hierarchical clustering and random permutation methods.

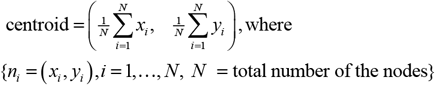

#### 2.1 Connectivity matrix of nodes from line segments

A hierarchical clustering algorithm requires distance metric between nodes. To meet the need, we developed a connectivity matrix using nodes from the line segments. We start by defining the first-order connectivity (**C**^1^) among the nodes. It is noteworthy that two nodes of a Hough line segment were trivially connected. We sought connectivity beyond such a trivial connection to have complete connectivity in the method described below. Specifically, we draw an extended line 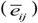 connecting arbitrary two nodes (*i*) and (*j*), and examine presence of any line segments that are found between two designated nodes or being in a proximity to 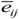 within tolerance levels based on angle and distance. In addition, we allowed gap between two nodes that any two nodes within a tolerance were marked as connected. Allowance of gap was necessary for enhanced connectivity between two nodes particularly located around corners because angles between the extended line 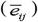 and nearby line segments were beyond a tolerance. In addition, we defined connectivity of a node to itself as zero. The first-order connectivity matrix is a square and symmetric matrix, which is an adjacency matrix with undirected edges in graph theory ^12^. The first-order connectivity was defined in Eq. (1). Details of construction of the first-order connectivity is presented in Supplementary Figure 6. The first-order connectivity matrix was then transformed into higher-order connectivity. Specifically, the second-order connectivity was defined as the result of multiplying the first-order connectivity matrix by itself, indicating the connectivity between two nodes through an intermediate node. Similarly, the *m*-th order connectivity (m > 1) was defined as the result of multiplying the first-order connectivity matrix m times, as shown in Eq. (2). We defined a total connectivity using connectivity matrix up to *m*-th order where *m* was set empirically to give entire connectivity of the nodes considered (*m* ≤5) in Eq. (3). The connectivity matrix was subsequently converted into a distance metric by taking the inverse of the exponentiated connectivity matrix, as described in Eq. (4).

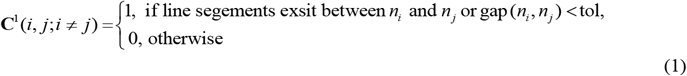

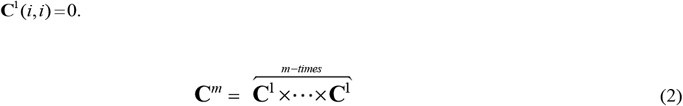

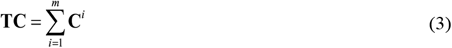

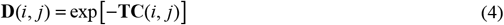

#### 2.2 Clustering of line segments to each pentagon

Using the distance matrix **D** in Eq. (4), we initiated clustering with the outer nodes. For the outer nodes, we applied a hierarchical clustering method using Wald’s minimum within-cluster variance criterion ^13^ and the distance metric described in Eq. (4). Additional details regarding the hierarchical clustering procedure can be found in Supplementary Material D. Following this step, each node was assigned a label indicating whether it belongs to pentagon 1 or pentagon 2.

To assign the inner nodes to their respective pentagons, we employed a random permutation approach in conjunction with hierarchical clustering. The inner nodes were initially grouped together based on their connectivity according to the distance matrix using hierarchical clustering. Subsequently, the classification of inner clusters into individual pentagons was achieved by randomly permuting the class labels (1 or 2) assigned to each inner cluster. The total number of random permutations, considering the number of inner clusters, increased exponentially. Each clustering result obtained with the inner nodes was paired with the clustering results obtained with the outer nodes to create a comprehensive clustering outcome. A diagram illustrating the combined clustering of outer and inner nodes is presented in Supplementary Figure 7.

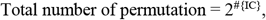

where #{IC} is the number of inner clusters. The total number of whole clustering results therefore is as much as the total number of permutations with the number of inner clusters.

#### 2.3 Determination of the best clustering outcome

Considering that a pentagon is a convex polygon, we aimed to reconstruct the entire PCT by combining two reconstructed individual pentagons. Each pentagon was reconstructed by applying a convex hull with the set of nodes assigned. We expected that the reconstructed PCT would exhibit minimal deviation from the original PCT if clustering of nodes to respective pentagon was done correctly. Therefore, the best clustering result was determined based on the minimum difference between the original image and the reconstructed image. To achieve this, we reconstructed a PCT by applying a convex hull to each of the comprehensive clustering results. The best clustering result was determined by evaluating the mean square error (MSE) of the residual image, which represents the deviation between the reconstructed PCT and the original image. Denoting the original image and the reconstructed image as I*o* and I*r*, respectively, MSE was calculated over a rectangle ? that covers both images, as defined in Eq. (5). A diagram illustrating this selection procedure was provided in Supplementary Figures 8 and 9. The best clustering result with a convex hull applied for each pentagon overlaid on original image is demonstrated in Figure 10.

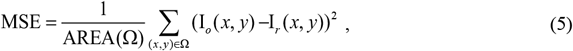

### Stage 3. Quantification of areas of interest

We quantify each area of pentagon using a built-in function of MATLAB (name: polyarea), which counts the number of pixels in a 2D polygon image. We quantify the areas of intersection using the Monte Carlo Integration ^11^, where we randomly draw samples (*x, y*) from a rectangle covering the intersecting area, and count the random samples falling inside the area of intersection as in Eq. (6). We denote the region of intersection of two pentagons as I_12._

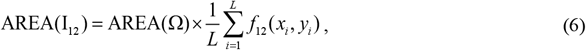

where (*x*_*i*_,*y*_*i*_), *i* = 1,2,…, *L*, are random samples selected from the inside of a rectangle ? that covers the entire PCT images I_1_ and I_2_., The function *f*_12_ is defined as

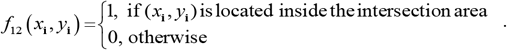

Using the measures of the areas of individual pentagons and their intersection, we quantify the total area of two interlocking pentagon, the proportion of the intersection as,

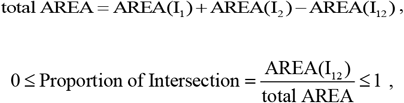

and a balance ratio of the two pentagon areas in terms of smaller area to larger area as follows.

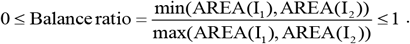

### Computation time

We measured the computation time using the sample pentagon, running on a 64-bit Linux system with a quad-core processor (with two threads per core) and 126 GB of random access memory. The computation time for the sample interlocking pentagon was 73 seconds. However, it’s worth noting that cases requiring manual corrections or exploration of non-default parameters set for the QIP which depends on the shapes of individual PCT may take longer time to process.

### Statistical Analysis

We adopted Python, R, and MATLAB at different stages and functions of the QIP algorithm. We fitted linear mixed effects model to longitudinal measurements of each quantification using age at baseline, sex, education, and lag (elapsed time at follow-up since baseline visit) as fixed covariates as well as subject intercept and slope as random effects to assess change over visits. The mixed effects model was executed using SAS 9.4.

## Data Availability

All data produced in the present study are available upon reasonable request to the authors.

https://www.radc.rush.edu

## Author contributions

NK and ST conceptualized and shaped the study’s framework and design. NK took the lead in designing the QIP algorithm, with contributions from TT. NK oversaw implementation of the QIP algorithm and conducted data analysis. NK drafted the manuscript. All authors (NK, TT, SDH, MH, ASB, DAB, and ST) participated in composition, review, and final approval of the manuscript.

## Data availability

Please visit the RADC Research Resource Sharing Hub (www.radc.rush.edu) to obtain data for research purposes.

## Figure legend

Figure 1. The diagram of the QIP algorithm. (a) Image of an interlocking pentagon, (b) Output of edge detection using the Canny edge detection algorithm, (c) Line segments detected through the Hough transformation, (d) Disentangled pentagons by clustering algorithms, and (e) Quantification of areas using the Monte Carlo integration.

Figure 2. Association among three area measures from the QIP algorithm. Pairwise associations were examined among three quantified measures: total area, proportion of intersecting area, and balance ratio. The associations were demonstrated as follows: (a) the association between proportion of intersecting area and total area, (b) the association between balance ratio and total area, and (c) the association between balance ratio and proportion of intersecting area.

Figure 3. Distribution of three metrics from the QIP algorithm. The distribution of the three measures (total area, proportion of intersection, and balance ratio) was compared among three age groups at baseline, categorized by age tertiles (age < 79, 79 ≤ age < 82, age ≥ 82).

Figure 4. Longitudinal patterns of three metrics from the QIP algorithm. Spaghetti plots of the three area measures derived from the QIP algorithm were presented as follows: (a) Total area, (b) Proportion of intersection area, and (c) Balance ratio.

## Supplementary Figures

**Supplementary Figure 1.**
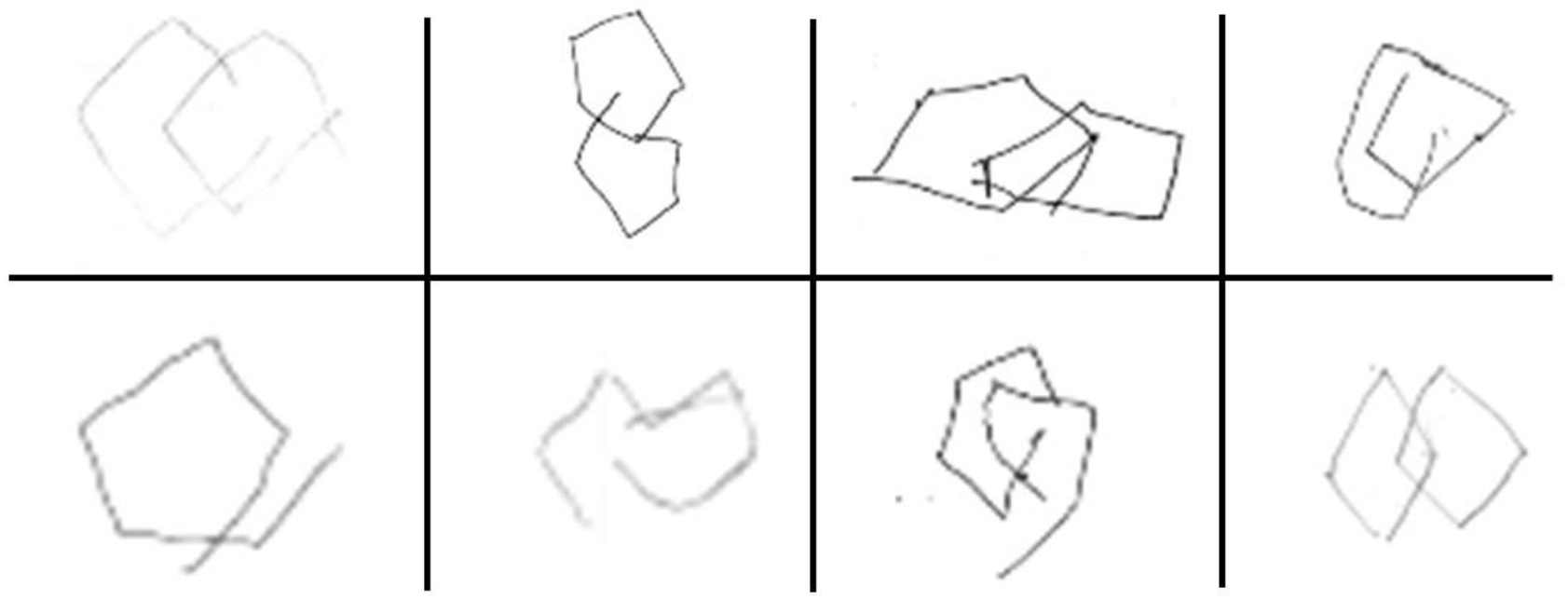
Examples of PCTs that failed the binary scoring criteria. Eight PCT images received a score of zero based on the binary scoring method.

**Supplementary Figure 2.**
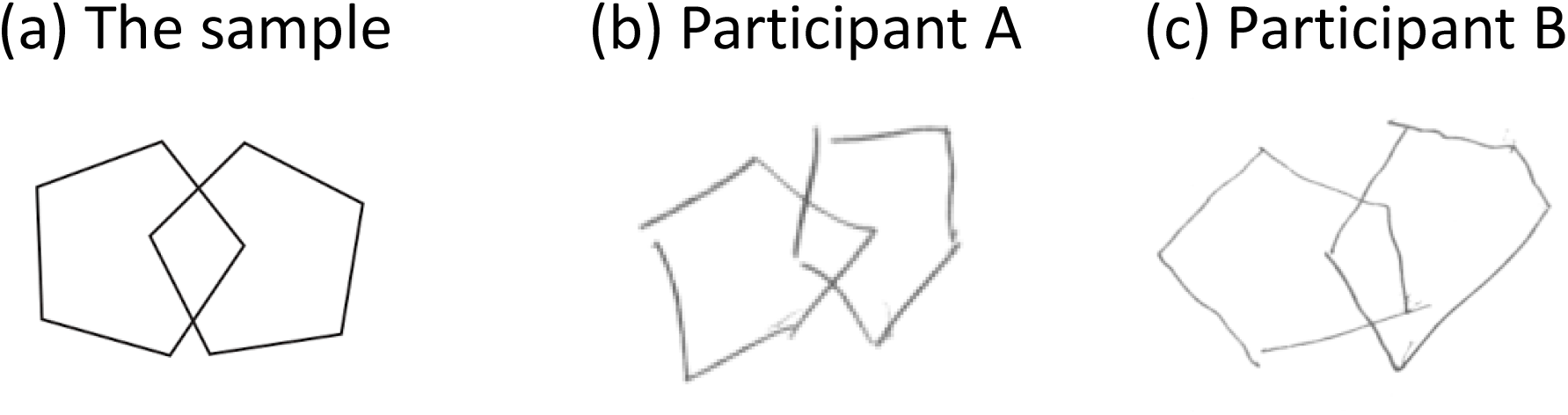
Three PCT images for demonstration. Three PCTs comprising the sample interlocking pentagon and two additional PCTs from different participants, whose ages ranged from their 70s to their 90s.

**Supplementary Figure 3.**
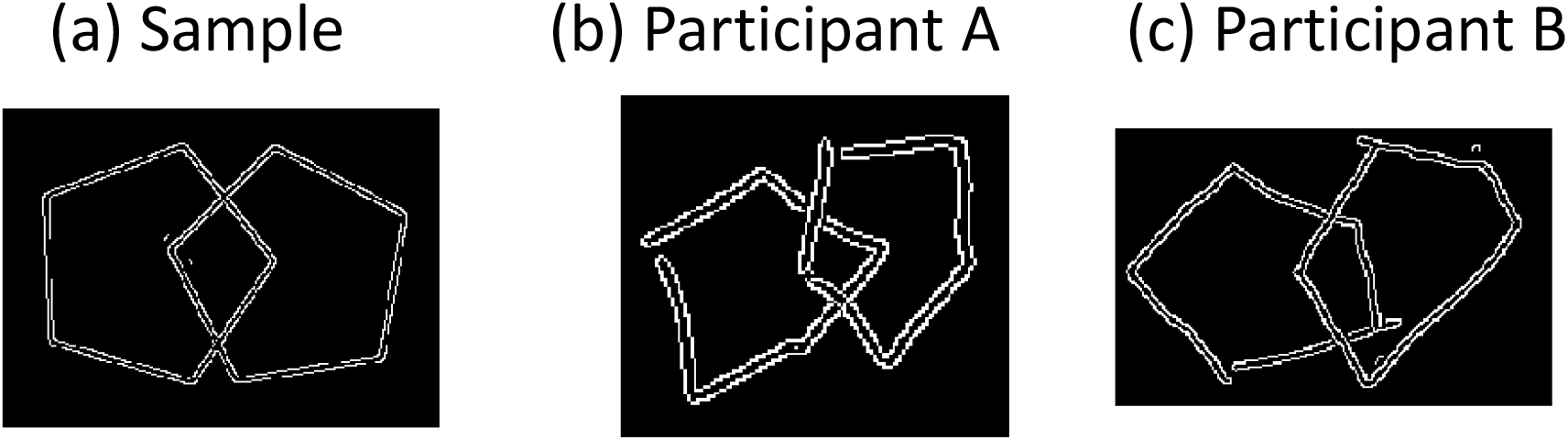
The Canny Edge Detection. The Canny edge detection method was applied to delineate the edges of each original PCT image shown in Supplementary Figure 2.

**Supplementary Figure 4.**
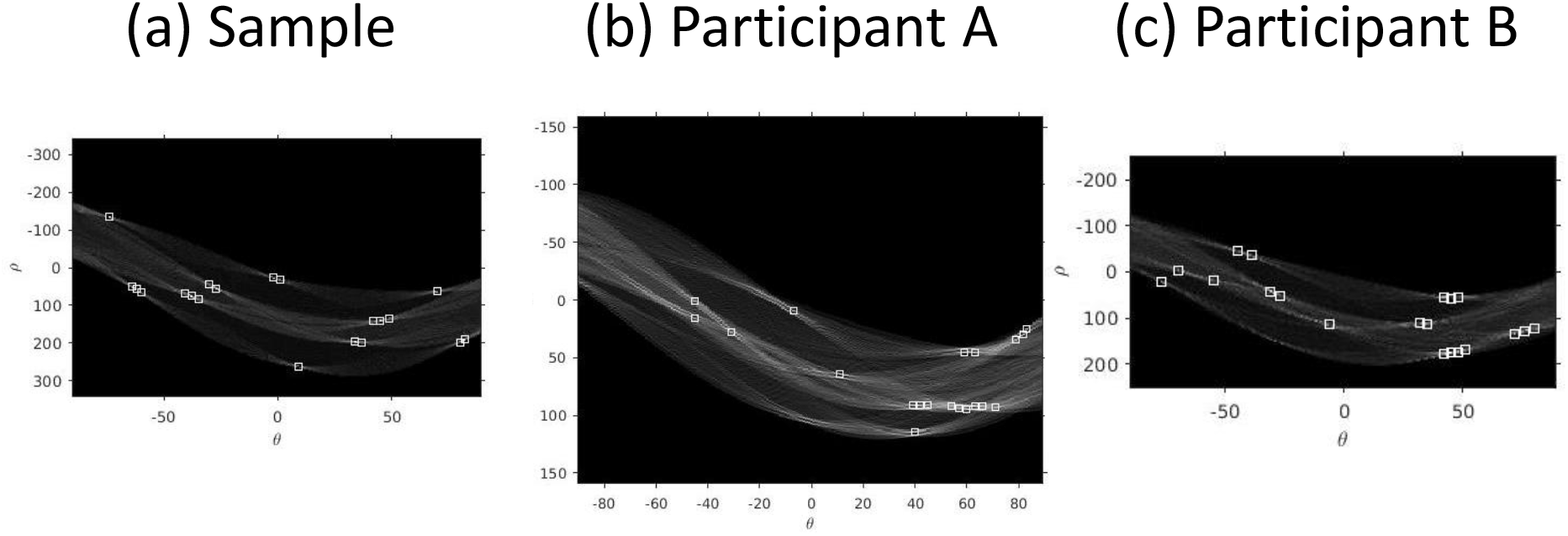
The Hough transformation. The Hough transformation with three demonstrated examples are represented graphically, where the x-axis corresponds to the angle (θ) between the perpendicular line to each line segment and the positive x-axis, and the y-axis represents the distance between the perpendicular line and the point (0, 0).

**Supplementary Figure 5.**
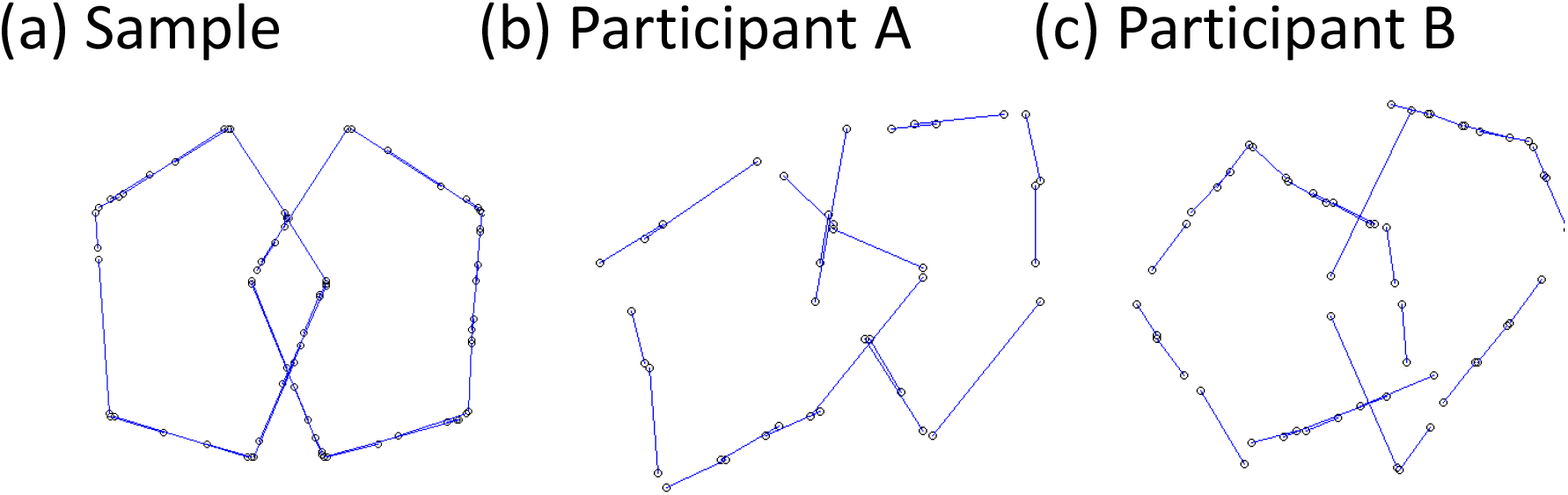
Line segments obtained from the Hough transformation. Line segments from the Hough transformation for three PCTs in Supplementary Figure2 were demonstrated.

**Supplementary Figure 6.**
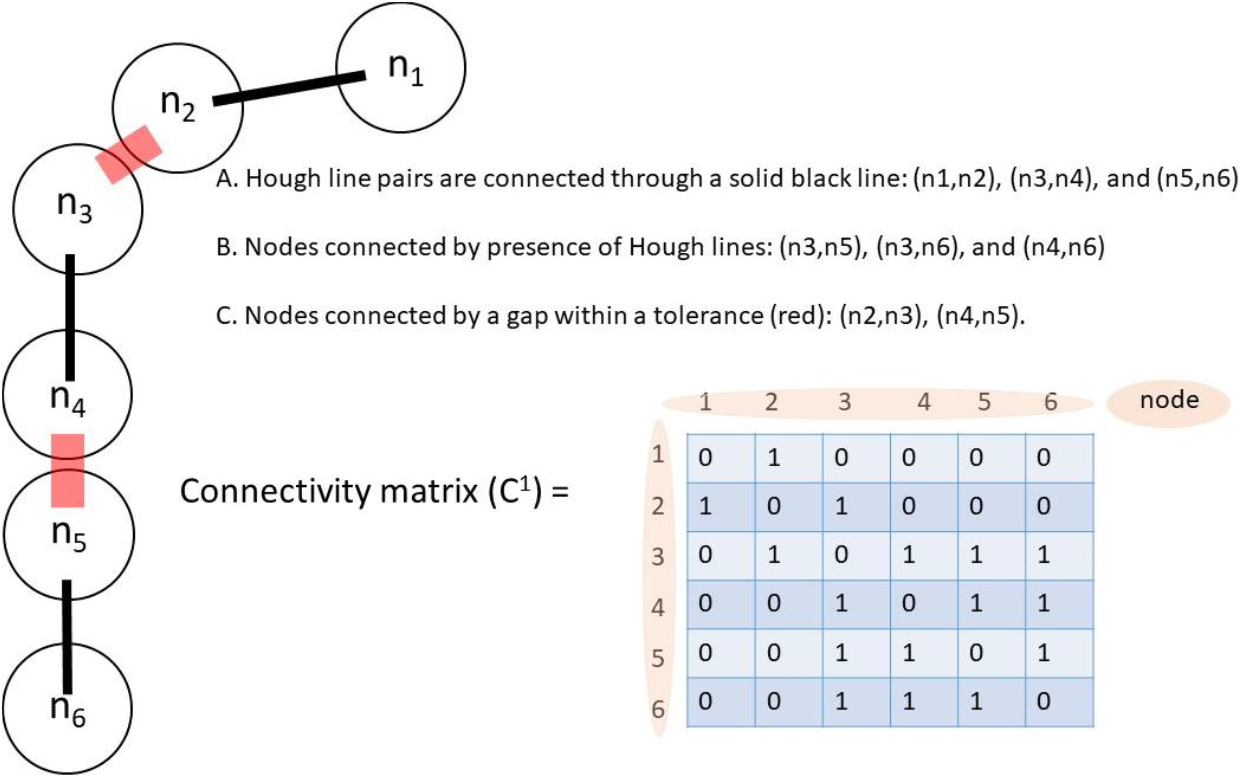
First-order Connectivity matrix. We demonstrated a first-order connectivity matrix consisting of six nodes.

**Supplementary Figure 7.**
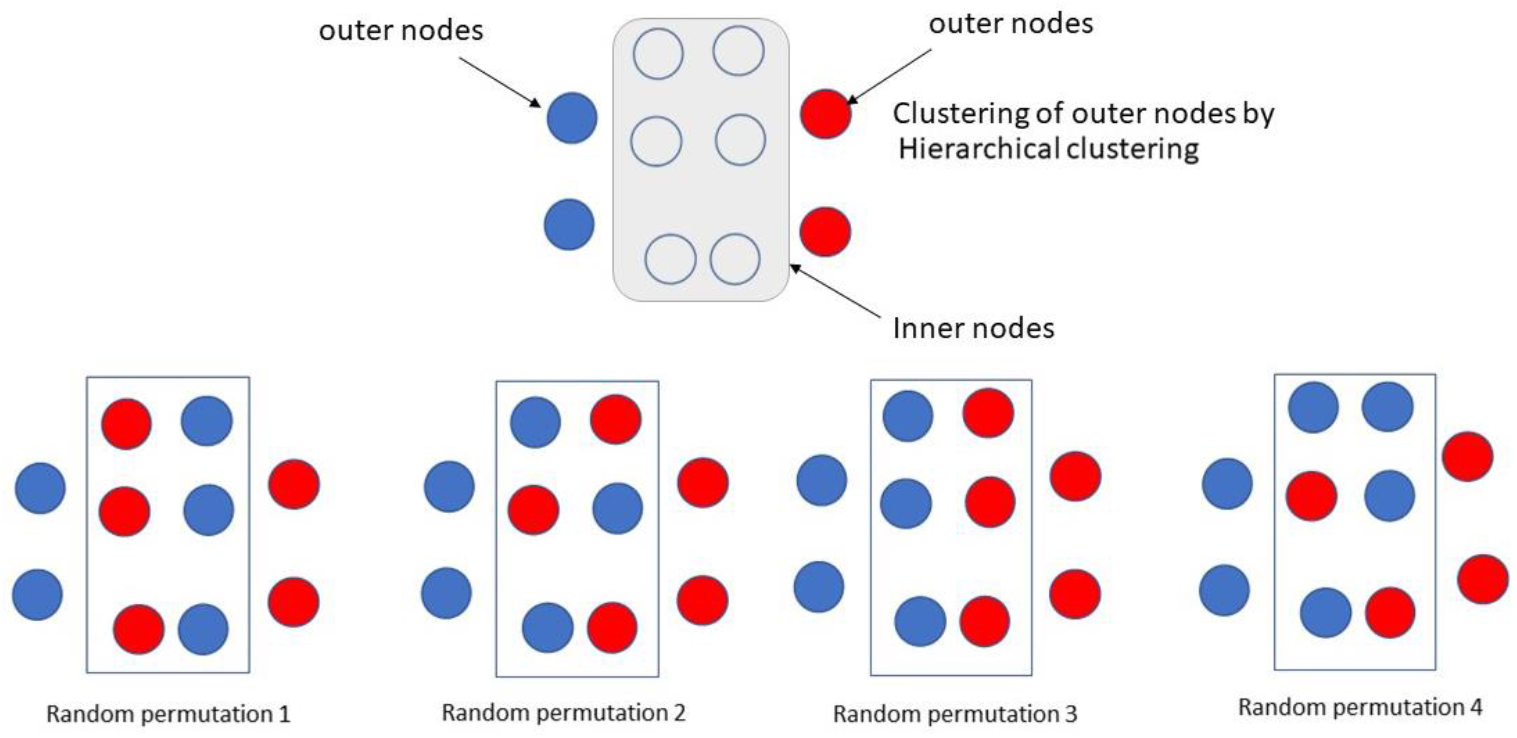
Clustering. A demonstration of complete clustering incorporating both outer and inner clustering results was performed. The outer nodes were clustered into their respective pentagons (red, blue). Subsequently, the inner nodes were clustered, and the resulting clusters were combined with the outer node clusters.

**Supplementary Figure 8.**
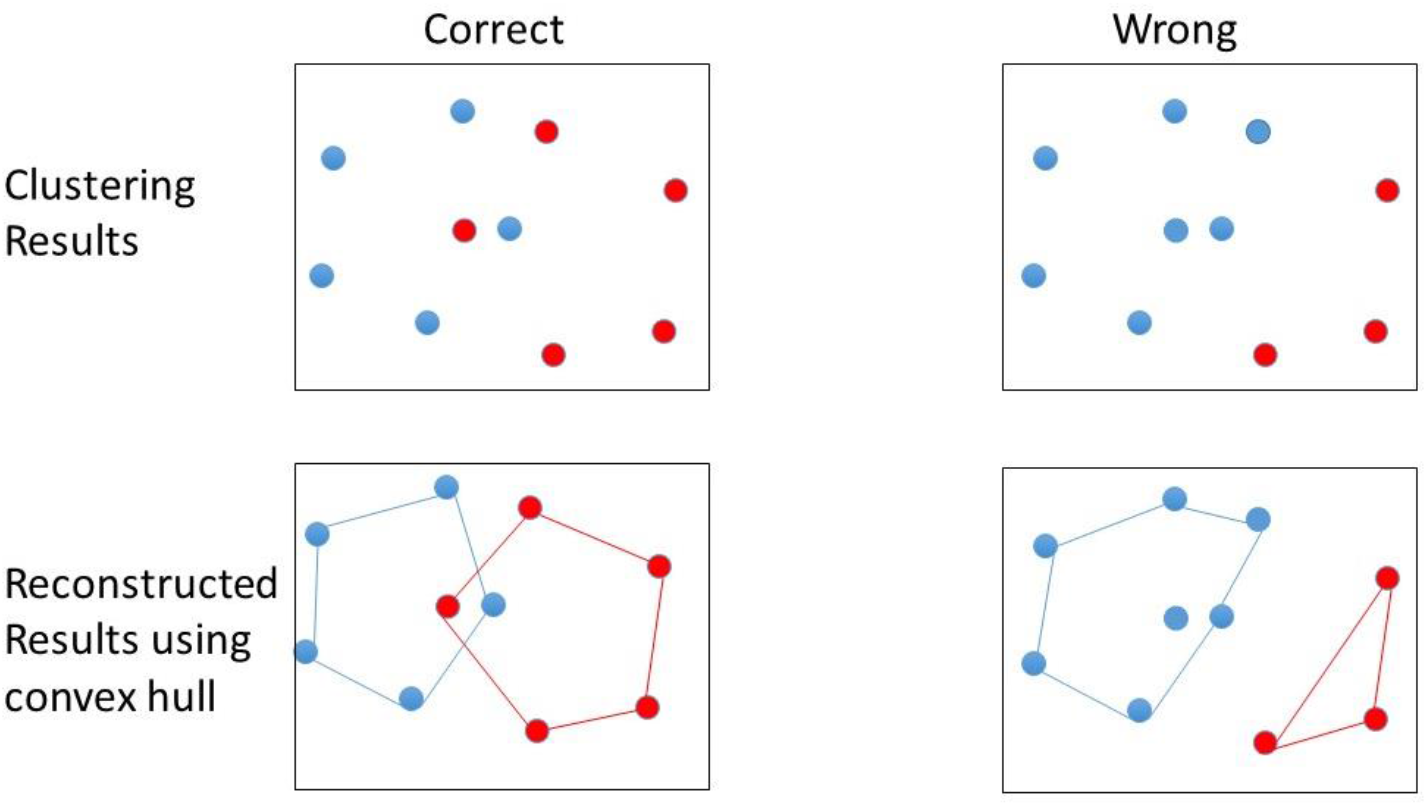
Reconstructed pentagons using convex hull after clustering. Demonstrations of the process showing reconstruction of pentagons by applying convex hull with clustering outputs.

**Supplementary Figure 9.**
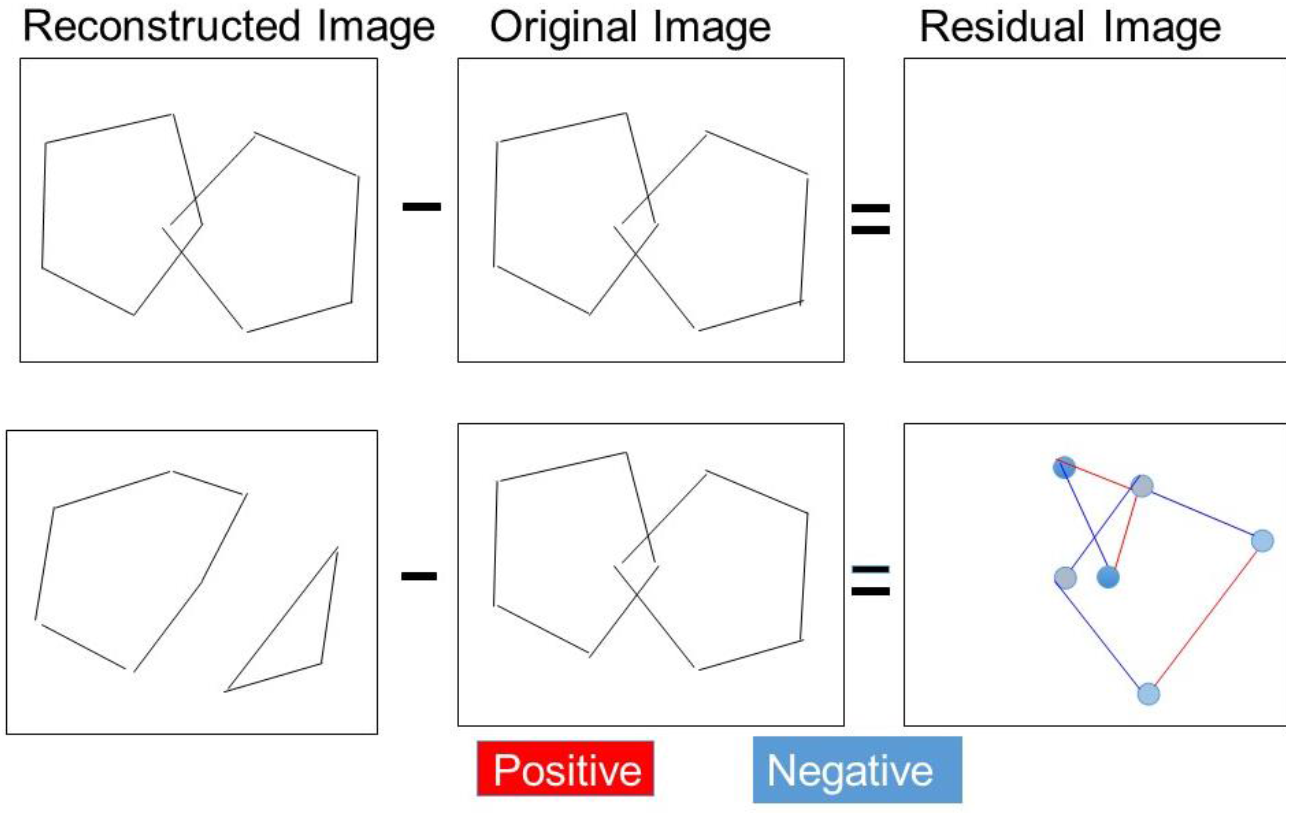
Residual images from the reconstructed and the original PCTs. The residual image was generated by subtracting the original PCT image from the reconstructed pentagons shown in Figure 10.

**Supplementary Figure 10.**
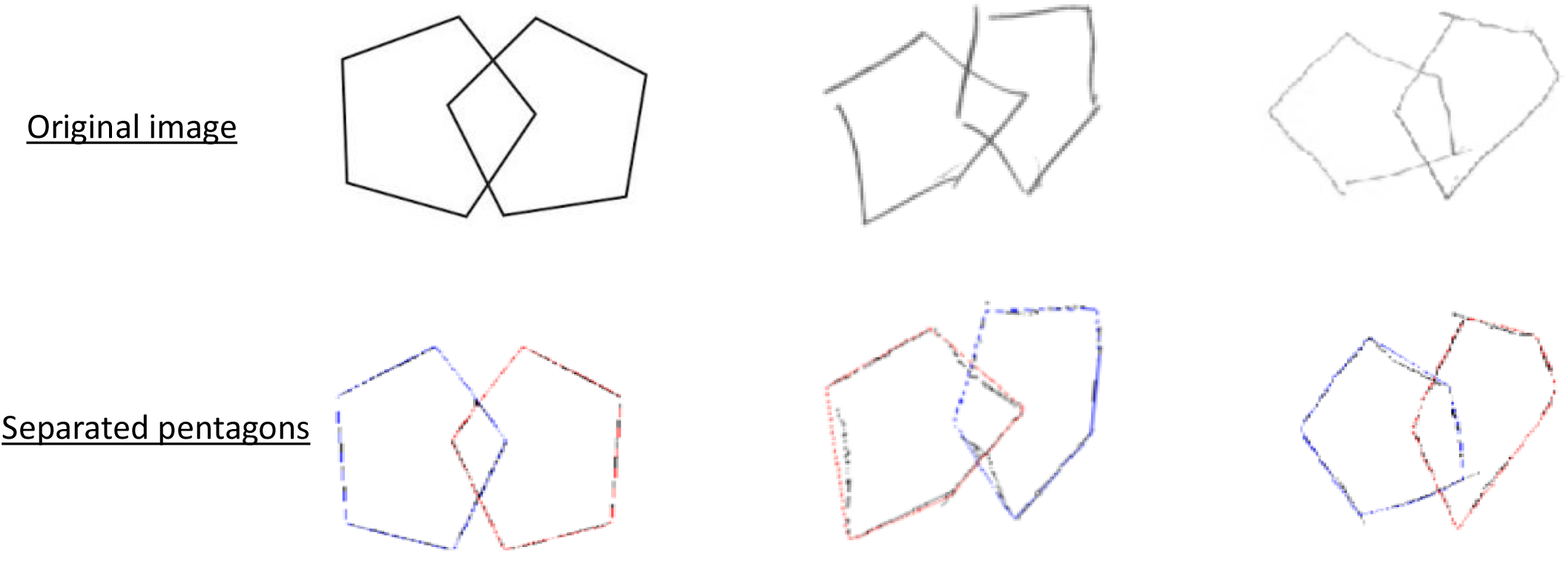
The best clustering results for the three demonstrated examples and the reconstructed pentagons using convex hull. The reconstructed pentagons, obtained using the convex hull with the best clustering result, were superimposed on each of the three presented examples.

**Supplementary Figure 11.**
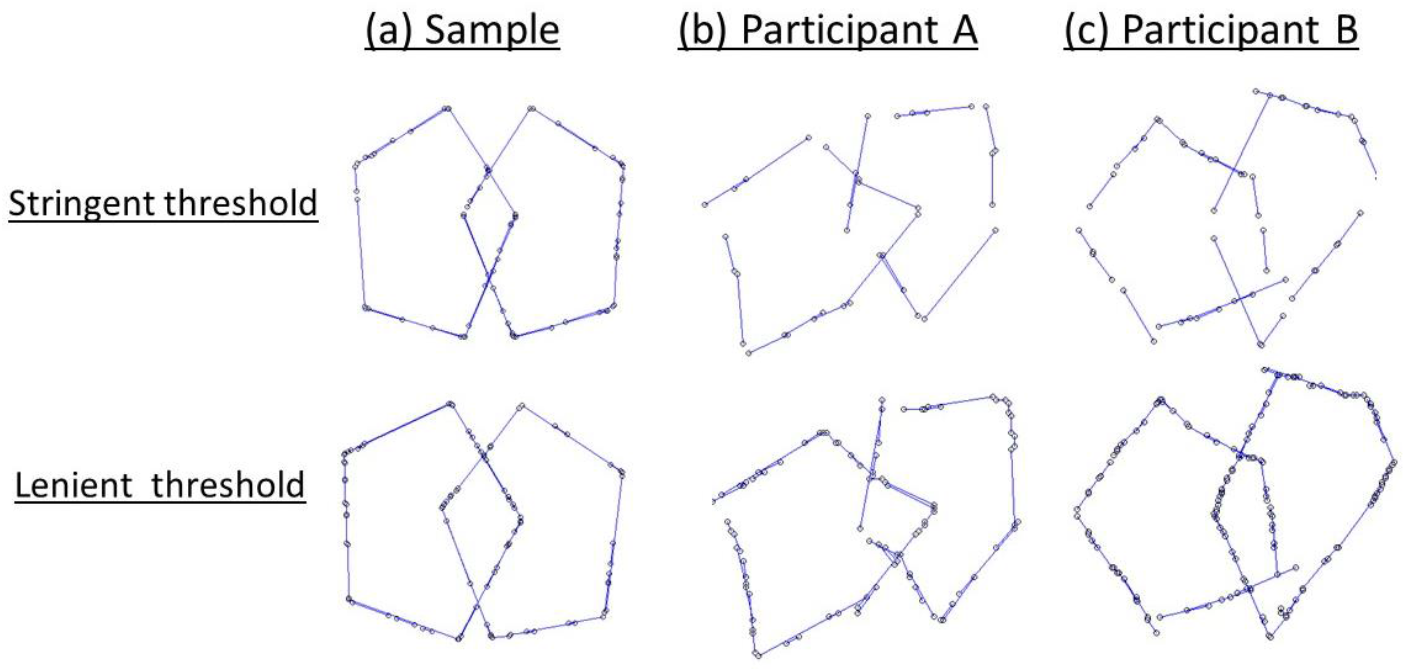
Line segments generated with two sets of thresholds from the Hough transformation. We present the differences in line segments using two sets of thresholds: stringent and lenient. The first row displays the line segments obtained using a set of stringent thresholds (10, 10, 4), while the second row shows the line segments obtained using a set of lenient thresholds (1, 2, 4). In these thresholds, the first component represents the minimal number of points sharing the Hough parameters, the second component denotes the minimal length of a line, and the third component indicates the minimal allowed gap between two distinct line segments.

