## Supplementary Materials for "A Three-Stage Algorithm for Quantification of the MMSE Interlocking Pentagon Areas"

### Supplementary Materials: Components of the QIP algorithm

#### A. The Canny edge detection

The Canny edge detection algorithm [9] simplifies the image by keeping only boundaries of the input image. Output from the Canny edge detection algorithm is a binary image with a positive value for detected edges. The Canny edge detection algorithm consists of five steps: (a) applying of a Gaussian filter to smooth the image; (b) assessing intensity gradient of the image using the Sobel filter; (c) suppressing non-maximum pixels to remove spurious response and thus make edges thinner; (d) applying double threshold to determine potential edges; and (e) edge tracking by hysteresis suppressing of all other edges that are weak or not connected to strong edges.

#### B. The Hough transformation

Images only with edges identified by the Canny edge detection algorithm [9] were transformed into the Hough space. Images were reconstructed with line segments identified from the Hough transformation [10]. To keep line segments with higher authenticity, three generally adopted thresholds were applied. Those are threshold ($\gamma_1$) for minimal number of points sharing the Hough parameters, a minimal length of a line ($\gamma_2$), and a minimal gap allowed between two distinct line segments ($\gamma_3$). Two Hough lines with a gap less than $\gamma_3$ were merged. More stringent thresholds remain only line segments with higher authenticity and longer length. However, applying higher thresholds could lose line segments that do not meet the thresholds such as short lines. We demonstrate such effects using two sets of thresholds, striengent vs. lenient, of $(\gamma_1, \gamma_2, \gamma_3)$. In Supplementary Figure 11, the first row shows the reconstructed images using a set of stringent thresholds (10, 10, 4), and the second row using a set of lenient thresholds (1, 2, 4). As results, reconstructed imges on the first row in the figure showed smaller number of line segments and longer length of the Hough lines with (10, 10, 4) than with (1, 2, 4). Furthermore, gaps between the Hough lines were larger with (10, 10, 4) because the lines that didn't meet the minimal number threshold ($\gamma_1$) and minimal length ($\gamma_2$) were discarded.

#### C. Designating nodes to inner and outer nodes

To distinguish between inner nodes and outer nodes, we introduced a cut-off distance from the centroid. Inner nodes were defined as nodes in close proximity to the centroid, while outer nodes were those located further away. To ensure optimal performance and avoid excessive processing time, it was crucial to select the appropriate set of inner nodes. To determine the set of inner nodes, we sorted the nodes based on their distance from the centroid in ascending order. Initially, we included nodes up to the 40th percentile as inner nodes. However, the cut-off distance could be adjusted based on the accuracy of clustering with the outer nodes. A higher cut-off distance might be necessary in cases where the quality of clustering with the outer nodes is compromised.

##### D. Hierarchical Clustering algorithm

Hierarchical clustering, employing Wald's minimum within-cluster variance criterion[13], is an agglomerative approach that constructs clusters by merging existing clusters at each step of the hierarchy. The objective is to minimize the within-cluster variance when determining which clusters to merge. Initially, each observation is considered as its own cluster at the first step and then grow combining other nodes closely connected.
